# New Insights on Excess Deaths and COVID-19

**DOI:** 10.1101/2020.07.24.20051508

**Authors:** Harry P. Wetzler, Herbert W. Cobb

**Affiliations:** Ofstead & Associates, Inc., 1360 Energy Park Drive, Suite 300, Saint Paul, MN 55108, USA; Independent Analyst, Coimbra, Portugal

## Abstract

**Background:** Weinberger and colleagues estimated that 27,065 of the 122,300 excess deaths in the United States between March 1 and May 30, 2020 did not have a COVID-19 cause of death.

**Methods:** The Centers for Disease Control and Prevention (CDC) post weekly data on mortality for 13 causes of death from the most prevalent comorbid conditions reported on death certificates where COVID-19 was listed as a cause of death. The 2015-2019 data for weeks 10 through 22 were used to forecast the number of deaths from the 13 causes in the absence of COVID-19 during 2020. The forecast was subtracted from the observed number of deaths for each cause during the period March 1 to May 30, 2020.

**Results:** The total of the differences for each of the 13 causes of death, 18,489 deaths, accounts for over two-thirds of the 27,065 excess deaths not due to COVID-19.

**Conclusion:** Combining the 95,235 reported COVID-19 deaths with the 18,489 from the 13 most frequent comorbid conditions reported on death certificates where COVID-19 was a cause suggests that as many as 93% of the excess deaths were due to COVID-19 and implies that COVID-19 deaths were undercounted. Ongoing assessment of excess deaths and causes of death is needed to provide a better understanding of the pandemic’s dynamics.

## Introduction

Weinberger and colleagues estimated that 27,065 of the 122,300 excess deaths between March 1 and May 30, 2020 in the United States (US) did not have a COVID-19 cause of death.^1^ The Centers for Disease Control and Prevention (CDC) post weekly data on mortality for 13 causes of death from the most prevalent comorbid conditions reported on death certificates where COVID-19 was listed as a cause of death. CDC states “Estimated numbers of deaths due to these other causes of death could represent misclassified COVID-19 deaths, or potentially could be indirectly related to COVID-19 (e.g., deaths from other causes occurring in the context of health care shortages or overburdened health care systems).”^2^ We used these data to provide insight on those 27,065 excess deaths with a cause of death other than COVID-19.

## Methods

The 2015-2019 data for weeks 10 through 22 were used in SPSS Forecasting to estimate the number of deaths from the 13 causes in the absence of COVID-19 during 2020.^3^ The forecast was subtracted from the observed number of deaths for each cause during the period March 1 to May 30, 2020. Confidence intervals were calculated using standard methods for sums and differences of random variables.^4^

## Results

The table shows the results using data submitted to CDC through June 13, 2020.

**Table.** Excess Deaths for 13 Causes of Death from March 1 to May 30, 2020.

**Table Excess Deaths for 13 Causes of Death from March 1 to May 30, 2020**
| **Underlying Cause of Death** | **2020 Forecast (95%CI)** | **2020 Observed** | **Difference** |
| --- | --- | --- | --- |
| Alzheimer disease and dementia | 69013 (65,014-73,012) | 77,621 | 8,608 |
| Cerebrovascular diseases | 38,429 (37,358-39,501) | 39,350 | 921 |
| Chronic lower respiratory disease | 41,693 (40,275-43,111) | 40,582 | -1,111 |
| Diabetes | 22,536 (21,571-23,502) | 25,480 | 2,944 |
| Heart failure | 23,152 (22,153-24,151) | 21,959 | -1,193 |
| Hypertensive diseases | 27,454 (25,745-29,163) | 29,630 | 2,176 |
| Influenza and pneumonia | 14,352 (13,241-15,462) | 17,080 | 2,728 |
| Ischemic heart disease (IHD) | 91,568 (90,568-92,569) | 96,309 | 4,741 |
| Malignant neoplasms | 148,120 (146,379-149,862) | 147,895 | -225 |
| Other diseases of circulatory system | 44,521 (43,445-45,597) | 43,018 | -1,503 |
| Other diseases of respiratory system | 17,428 (16,632-18,223) | 17,479 | 51 |
| Renal failure | 12,543 (12,019-13,067) | 12,901 | 358 |
| Sepsis | 10,052 (9,334-10,769) | 10,046 | -6 |
| **Totals** | **560,861** | **579,350** | **18,489** |

The total difference listed above, 18,489 (95% confidence interval: 12,652 - 24,326) deaths, accounts for over two-thirds of the 27,065 excess deaths not due to COVID-19 and leaves 8,576 deaths due to other causes.

## Discussion

Combining the 95,235 reported COVID-19 deaths with the 18,489 from the 13 most frequent comorbid conditions reported on death certificates where COVID-19 was a cause suggests that potentially 93% of the total excess deaths were due to COVID-19. Moreover, some of the 8,576 deaths due to other causes may have been COVID-19 deaths. The 93% value is consistent with an estimate of 95% of excess deaths attributed to COVID-19 in the US in April 2020.^5^ Compromised care processes (e.g., lack of intensive care capacity) combined with incomplete COVID-19 testing could decrease the attribution of COVID-19 as the cause of death.

Furthermore, the unknown manifestations and pathology of COVID-19 including being asymptomatic are additional factors for missed COVID-19 as the cause of death. Based on the excess death data and known issues in the process of care and identification, we submit that most of these deaths were due to COVID-19. For instance, the difference for Alzheimer disease and dementia is likely due in large part to long term care deaths that were actually COVID-19 related. The difference for IHD may be due to deaths where care was not sought due to fear of COVID-19 or from suboptimal care due to COVID-19 overload.^6^ Diabetes and hypertensive diseases have been widely mentioned as increasing COVID-19 mortality risk and those deaths may actually have been caused by COVID-19. The increase in influenza and pneumonia deaths may reflect instances where virus testing was not available.^7^ The negative results for chronic lower respiratory disease, heart failure, and other diseases of the circulatory system may reflect situations where COVID-19 pre-empted these causes.

## Conclusion

Combining the 95,235 reported COVID-19 deaths with the 18,489 from the 13 most frequent comorbid conditions reported on death certificates where COVID-19 was a cause suggests that as many as 93% of the excess deaths between March 1 and May 30, 2020 were due to COVID-19 and implies that COVID-19 deaths were undercounted. Ongoing assessment of excess deaths and causes of death is needed to provide a better understanding of the pandemic’s dynamics.

## Data Availability

Data used in this study are available at: https://data.cdc.gov/NCHS/Weekly-counts-of-death-by-jurisdiction-and-cause-o/u6jv-9ijr

